# Autistic Psychiatrists’ Perspectives on Mental Healthcare for Autistic People: A Qualitative Study

**DOI:** 10.64898/2026.06.01.26354595

**Authors:** Mary Doherty, Nicholas Chown, Nicola Martin, Bernadette Grosjean (Ret), Eddie Chaplin, Sebastian C. K. Shaw

## Abstract

**Background:** Autistic people experience disproportionately high rates of co-occurring mental illness and suicide, yet mental healthcare services routinely fail to meet their needs. Patients unrecognised as autistic are at risk of ineffective or harmful treatment. Autistic psychiatrists occupy a unique position: as members of both medical and autistic communities, they offer dual insider perspectives that may directly shape patient outcomes. Despite being the second largest specialty group in Autistic Doctors International (ADI), this workforce remains largely unrecognised and underutilised. This study examines autistic psychiatrists’ perspectives on mental healthcare for autistic people.

**Methods:** Loosely structured interviews were conducted with seven senior autistic psychiatrists across child and adolescent, adult, and liaison psychiatry, recruited from a psychiatry-specific subgroup of ADI. Data were analysed using reflexive thematic analysis: codes related to patient care and mental health services were extracted and analysed as a focused subset.

**Outcomes:** Nine themes were identified: autistic-to-autistic therapeutic rapport; benefit of recognition and diagnosis; early recognition and education as preventive factors; iatrogenic harm from non-recognition and systemic pathways to misdiagnosis; knowledge gaps and stereotypes; inaccessible services; resource constraints and diagnostic thresholds; autistic psychiatrists as an underutilised resource; and pathways to change.

**Interpretation:** Autistic psychiatrists’ dual insider positionality affords a unique and under-acknowledged vantage point on what autistic patients experience and where mental healthcare fails them. The mental health burden autistic people carry is substantially shaped by systems not designed for them. Embedding neurodiversity-affirmative practice, closing training gaps, reforming diagnostic pathways, and recognising autistic psychiatrists as a clinical and epistemic resource offer a coherent pathway to improving mental health outcomes for autistic people.

**Funding:** None

**Research in context:** *Evidence before this study:* Autistic people experience disproportionately high rates of co-occurring mental illness and suicide, yet mental health services consistently fail to meet their needs. Existing literature documents barriers to care, inadequate training across psychiatric curricula, and structural failures including inaccessible crisis services and poorly configured diagnostic pathways. Qualitative research has explored autistic adults’ experiences of receiving mental healthcare, and participatory and insider methodologies in autism research have yielded insights inaccessible to outsider researchers. A previously published qualitative study of autistic psychiatrists, from which the data for the current study are drawn, examined their experiences of recognising themselves and their colleagues as autistic, and identified dual insider positionality as a distinctive feature of their professional experience. Notably, participants described retrospectively recognising that, before their own autistic identity had been acknowledged, they had missed autistic patients; their unrecognised neurodivergence skewing the clinical yardstick against which they assessed others. However, no previous study has examined what autistic psychiatrists observe from this dual insider position in relation to patient care: what works, what causes harm, and what needs to change.

*Added value of this study:* This study is the first to examine autistic psychiatrists’ patient-facing perspectives using insider qualitative methodology. Drawing on data from seven senior autistic psychiatrists across multiple UK specialties, it documents nine themes spanning therapeutic rapport, the transformative benefit of neurodevelopmental recognition, preventable harm when recognition is absent, structural and attitudinal barriers to care, and the daily clinical reality of resource constraints and diagnostic thresholds. The insider perspective reveals both what effective autistic–autistic clinical encounters produce and the systemic conditions that prevent their benefits from reaching the majority of autistic patients. Crucially, participants articulate the tension between neurodiversity-affirmative values and resource-rationed systems, grounding the analysis in clinical reality rather than abstract aspiration.

*Implications of all the available evidence:* The combined evidence identifies autistic psychiatrists as a largely unrecognised clinical and epistemic resource whose distinctive expertise – in recognition, rapport, and systemic understanding – should inform training, workforce planning, and service development. Curricular reform with training delivered by autistic clinicians, diagnostic pathway redesign to remove structural incentives for misdiagnosis, and neurodiversity-affirmative approaches embedded in education from the earliest opportunity represent achievable, evidence-informed changes with potential to substantially reduce the preventable mental health burden autistic people carry.

## Introduction

Autistic people experience disproportionately high rates of co-occurring mental illness, including anxiety, depression, and post-traumatic stress disorder, with suicide rates multiple times that of the general population.^1,2^ Mental health services are frequently cited as failing to meet autistic people’s needs: autistic patients report difficulty feeling understood by clinicians, describe misinterpretation of their behaviour and communication as barriers to engagement, and face elevated risk of receiving ineffective or harmful treatment when their neurodivergence is unrecognised.^3^ Diagnostic pathways are poorly configured for autistic presentations, reasonable adjustments are under-implemented, and the transition between services represents a period of particular vulnerability.^4–7^

The drivers of these failures are increasingly documented. Training in autism across medical and psychiatric curricula remains inadequate, relying on outdated stereotypes that systematically exclude women, people from minoritised ethnic groups, and those who have developed effective masking strategies.^8–11^ Milton’s double empathy problem^12^ – the bidirectional difficulty in mutual understanding between autistic and non-autistic people – has implications for clinical practice: therapeutic relationships, diagnostic processes, and service design have been developed predominantly by and for neurotypical people, with autistic experience treated as deviation from a non-autistic norm.^13^ A neurodiversity-affirmative model, which frames autism as a form of natural human variation rather than deficit, has been proposed as a more appropriate basis for clinical care, but its uptake in mental health services has been uneven.^14,15^

Lived experience perspectives have been increasingly recognised as essential to understanding and improving healthcare for marginalised groups.^16^ Within autism research, participatory and insider methodologies have yielded insights inaccessible to outsider researchers, including the reframing of diagnostic criteria, the identification of iatrogenic harm, and the articulation of service priorities from an autistic standpoint.13, 17, 18 Autistic healthcare professionals occupy a particularly distinctive position: as members of both the medical profession and the autistic community, they possess dual insider positionality – simultaneous access to clinical knowledge and autistic lived experience – that may afford unique insight into where and why healthcare systems fail autistic people.^13,19^

Autistic doctors are present across medical specialties; psychiatrists represent the second largest specialty group within Autistic Doctors International (ADI), a global network of over 1,500 autistic medical professionals.^20^ Existing research has examined autistic doctors’ own experiences of working in medicine, including challenges of disclosure, professional identity, and workplace adjustment.^21,22^ However, the perspectives of autistic psychiatrists on mental healthcare for autistic people – what works, what causes harm, and what needs to change – have not been systematically examined. This represents a significant gap: the clinicians best positioned to identify both what good care looks like and where the system is failing remain an underutilised source of knowledge.^23^

This paper examines the perspectives of senior autistic psychiatrists on mental healthcare for autistic people, drawing on qualitative interview data to explore what their dual insider positionality affords in understanding both effective care and systemic failure.

## Methods

### Design and ethical approval

This study reports a focused subset analysis of qualitative interview data from a previously published study of autistic psychiatrists’ experiences of recognising themselves and others as autistic.^20^ Full details of study design, ethical approval (LSBU ETH2122-0128), recruitment, and data collection are reported in the primary publication. Briefly, loosely structured interviews were conducted with senior autistic psychiatrists recruited through purposive sampling from a psychiatry-specific subgroup of Autistic Doctors International. Eight participants were initially recruited; one was excluded following institutional review due to concerns about identification. The analysis reported here is based on data from the remaining seven participants.

### Participants

Seven senior UK-based autistic psychiatrists participated (Table 1). Specialties included child and adolescent, adult, intellectual disability, old age, and liaison psychiatry. Five held consultant posts; two were senior specialty doctors. Six were in current clinical practice; one was retired. One participant held a formal autism diagnosis received in adulthood; one had a childhood PDD-NOS diagnosis and self-identified as autistic as an adult; five self-identified as autistic.

**Table 1:**
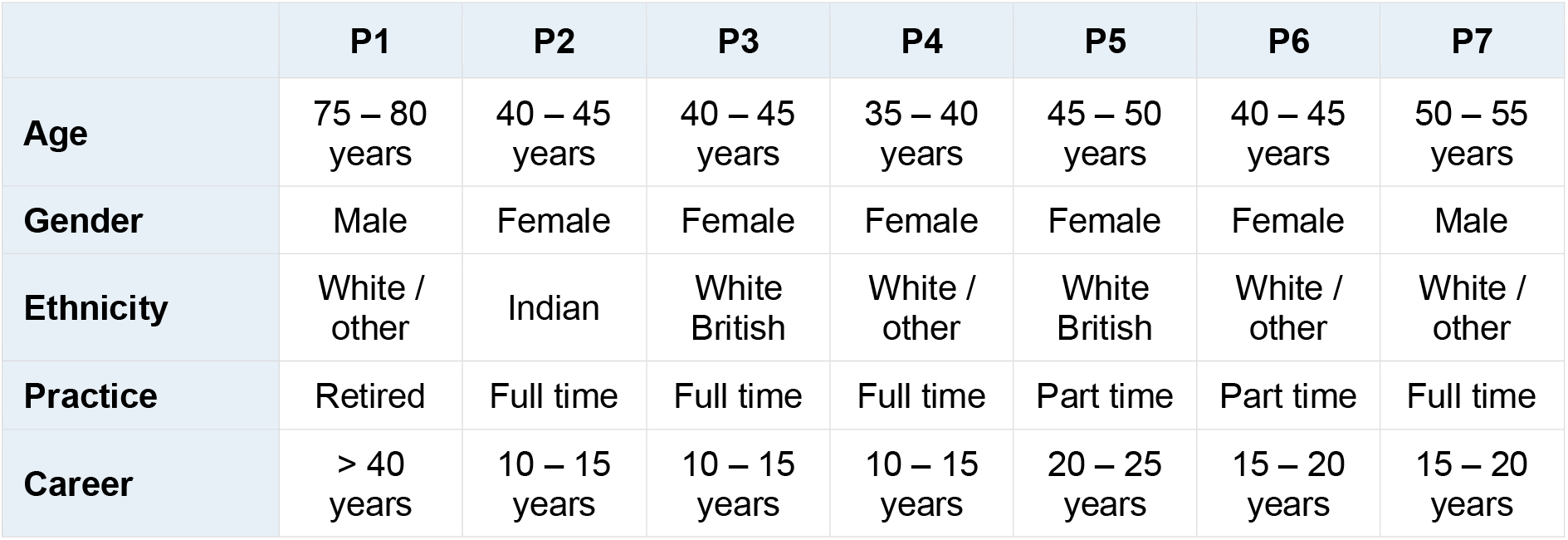
Participant Demographics.

|  | P1 | P2 | P3 | P4 | P5 | P6 | P7 |
| --- | --- | --- | --- | --- | --- | --- | --- |
| **Age** | 75 – 80 years | 40 – 45 years | 40 – 45 years | 35 – 40 years | 45 – 50 years | 40 – 45 years | 50 – 55 years |
| **Gender** | Male | Female | Female | Female | Female | Female | Male |
| **Ethnicity** | White / other | Indian | White British | White / other | White British | White / other | White / other |
| **Practice** | Retired | Full time | Full time | Full time | Part time | Part time | Full time |
| **Career** | > 40 years | 10 – 15 years | 10 – 15 years | 10 – 15 years | 20 – 25 years | 15 – 20 years | 15 – 20 years |

### Analysis

Interview transcripts were re-analysed using reflexive thematic analysis, with codes relevant to mental healthcare for autistic people extracted as a focused subset for concentrated re-analysis, involving iterative coding, reflexive journaling, and collaborative theme development.^24,25^ The first author’s positionality as an openly autistic clinician and insider researcher facilitated interpretive depth and access to meaning that may have been inaccessible to outsider researchers. Analytic rigour was maintained through reflexive journaling throughout the analytic process, iterative challenge between the two co-analysts (MD, an autistic clinician, and NC, an autistic academic), and sustained attention to data that complicated or resisted straightforward interpretation. Themes were developed inductively from the data and refined through multiple analytic cycles.

### Positionality statement

The research team is autistic-led; the first author is an openly autistic clinician and insider researcher, with this dual positionality informing reflexive practice throughout the analysis.^19^ Three further co-authors – NC, BG and SCKS – also identify as autistic. BG is a psychiatrist.

## Results

Analysis resulted in nine distinct themes (Figure 1). Our results include one to two anchor quotes per theme, with a wider range of evidencing quotes which are sequenced thematically in Table 2.

**Figure 1.**
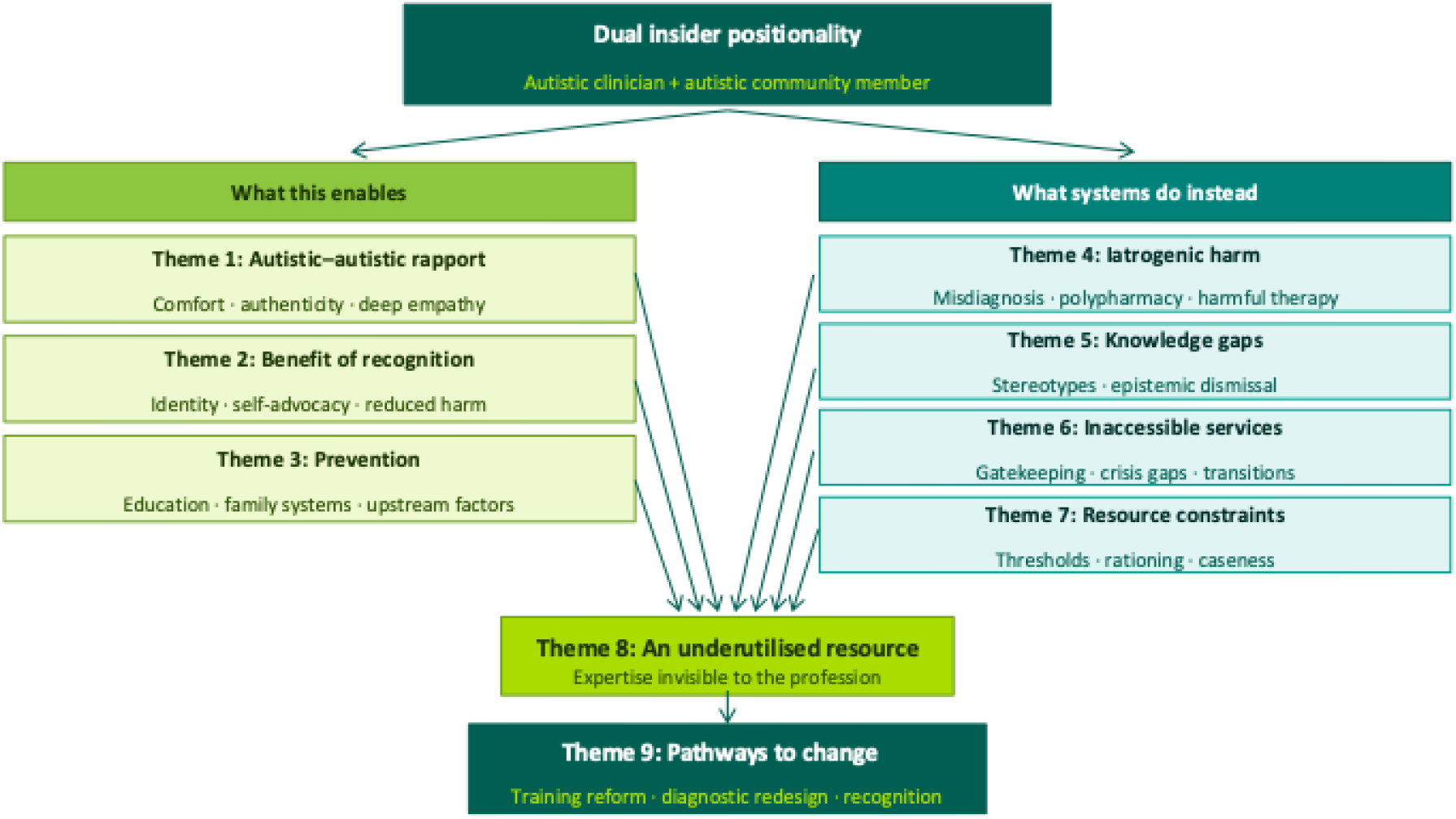
Dual insider positionality as a clinical and epistemic resource: A conceptual model of nine themes from autistic psychiatrists’ perspectives on mental healthcare. Dual insider positionality – simultaneous membership of the clinical profession and the autistic community – generates both distinctive clinical capabilities (left) and insight into where systems fail (right), resulting in an underutilised workforce resource with clear implications for change.

**Table 2:** Illustrative Quotes by Theme.

| Theme | P# | Illustrative Quote |
| --- | --- | --- |
| **Theme 1: Rapport** | P3 | “A couple of people come to mind where they said ‘Oh no one really understood, no one understood this side of things” |
|  | P3 | “I’ll give examples without realising, really specific examples... they’ve given me a knowing look, nothing more, nothing actually said explicitly... moments of shared understanding definitely” |
|  | P4 | “They come with their neurotypical parents... the parent is like ‘I’m so sorry, he’s going on about it and it makes no sense’... I was like ‘it makes perfect sense’... the parents are like... ‘what the hell has just happened” |
|  | P4 | “I have a lot of neurodiverse kids on my caseload... they send them to me when everybody else can’t do anything or they get frustrated with them so I have kind of accumulated a nice flock” |
|  | P4 | “Somebody who can translate... from neurodivergent to neurotypical, being that person in the middle who speaks both languages and can explain to the parents in the way that they get it” |
|  | P4 | “Kids absolutely love it... they’re like oh look at all those fidget toys and squeegies and some of them I just can’t get them out of my sensory cave” |
|  | P6 | “There’s been a couple of times with people where they’ll be saying things [about] being autistic and I might kind of wink at them and go ‘yeah well who’d wanna be neurotypical anyway’... that kind of lets them know I’m onside or might have them wondering about me” |
|  | P7 | “Some patients say you are too aren’t you... I say well possibly yes, never diagnosed but maybe” |
|  | P1 | “If I’m dealing with an issue where autism is central it’s very difficult not to have my views on my autism influence it and so I’ve got to try and stand back and look at their autism... it can be very unhelpful and if anything sidetrack it to talk about my autism and your autism and it did get just too complicated” |
|  | P2 | “I’m not open, I’ve never told any of my young people that I am too... what I do tell them is... whatever I know is... not from my textbooks, it’s from lived experiences but not specifically my own... I wish I could though because it is mostly my own” |
| **Theme 2:**<br>**Recognition** | P5 | "The biggest difference to their life that any doctor had ever made and I did not even say the word autism" |
|  | P3 | "It's the understanding that's the number one thing that's important more than anything else because [if] you totally understand yourself you can advocate I think much better for yourself if you totally understand where you come from and been brought up with a positive sense of who you are and what you need" |
|  | P7 | "We ask the questions... start the process to get them off some of the medication and helping them better and... obviously if people don't need to be on ridiculous amounts of medication for presumptive mood disorders or psychosis then that's a very good outcome for them" |
|  | P4 | "We can modify the environment, get the person a bit of empowering advice and off they go... you have them off your caseload straight away" |
|  | P2 | "I don't use the word autism, I say neurodiversity... because I'm not allowed to hand out a diagnosis... it does seem to help a lot" |
|  | P2 | "I demonstrated it very easily because I know what questions to ask... if I empty the streets of human beings would you go out... and they are very happy to go out then" |
|  | P6 | "I thread my formulation with the most minutest of details and the most oddest of funny stories... I will use those to literally pull the whole thing together like I'm weaving tapestry with tiny cotton" |
|  | P5 | "[A] person... coming to the end of their life... very obviously autistic to me... sort of broached it in a sideways manner... feeling a bit different... in the family...and although we never got as far as diagnosing them... they clearly found some kind of comfort from knowing before they died" |
| **Theme 3:**<br>**Prevention** | P2 | "If they have not been seeing a psychiatrist for years... the suicidality and self-harm seems to melt away... but if they have been on our caseload for years without a diagnosis it's quite entrenched by the time they reach me" |
|  | P3 | "They've let her go in early before everybody else... come in five minutes early to be away from all the noise and the rush... they've given the cards on the desk and they know and they accept that she's autistic without a diagnosis... we don't need an EHCP and they're putting in the smaller scale interventions" |
|  | P4 | "No energy left for learning if all used up trying to comply with behavioural expectations" |
|  | P4 | "We have sent quite a few parents to the adult service and they have now been diagnosed and medicated and that has been the best intervention for the family... we have nice success stories of... parents getting assessed, getting some treatment and then the whole family kind of thriving a bit more because the parents are getting the right help for the ADHD and the social worker is not on the case all the time for being a bad parent" |
|  | P7 | "[Diagnosis] can help but it doesn't take away the low mood which they've typically usually had since their teens... I would like to think that people would be picked up earlier for starters" |
|  | P5 | "Had I known as a child I think that the medical model would have been very much applied, I would have been directed out of medicine into something... that didn't match my skills and strengths and my intense |
|  |  | interest... would that have given me half a life and made me exceedingly depressed... and everybody would have thought well that's just autistic people, that's just what happens, it's part of it – which I really don't think is the case for most of us” |
|  | P6 | “Building that strength-based narrative... I think it would... help patients... view it as something actually that could be a positive thing” |
| **Theme 4:**<br>**iatrogenic harm** | P4 | “All this kind of iatrogenic harm that we sometimes do because you haven't looked at the beginning through the neurodevelopmental lens” |
|  | P4 | “I'm 90% certain already, from what I have heard, this is an autistic person... really struggling and suffering... they can't make themselves any different and yet you're just gonna put more layers of punishment” |
|  | P2 | “The adult wards are full of EUPD people... EUPD like autism is seen as something that is just blacklisted... nothing can be done... the teams are becoming more neurodivergent aware but still...” |
|  | P7 | “Wherever I go and acquire caseload I find people who've been on there for years often with varying or vague diagnoses and polypharmacy who are blatantly neurodevelopmental and nobody asked the question” |
|  | P2 | “A lifetime diagnosis of bipolar or borderline and treatment resistant depression and anxieties... but you don't hear the developmental aspect of it easily so unless... somebody has put in the effort to go into it” |
|  | P3 | “I guess it's much easier isn't it to put a BPD diagnosis on somebody” |
|  | P5 | “If you're more confident making those other diagnoses, you're going to spot the features more of those, aren't you” |
|  | P1 | “When I was training as child psychiatrist I'd be seeing these children, adolescents, who'd got autism and intellectual disability, saying no they're not autistic it's intellectual disability... and then... seeing the same children three years older and saying our thinking has shifted a bit now, really sorry, we got this wrong and you're quite right he's autistic” |
|  | P4 | “The waiting list and the hoops and everything... this is the only diagnosis where this is required... if I wanted to diagnose bipolar, schizophrenia whatever, I take as long as I think it's necessary, go to the criteria and make the diagnosis, I could do it in 5 minutes if I was so inclined... there isn't that kind of prescriptive... pathway... why is it different for autism? It seems to create this big barrier for actually picking people up and I do wonder whether it creates some pervasive incentive to actually use personality disorder instead because you don't have to go through the hassle of referring to specialist service and doing all this stuff... if you are doing it well you should have psychological assessments and scales and I suppose probably 90% of people don't bother... is there something in the system that actually pushes people away from neurodevelopmental to personality just for like speed and ease... people are people, people want easy life... that's generally how human species operates” |
| **Theme 5:**<br>**Knowledge gaps** | P3 | “The one thing I'm most sure about is she's not autistic... she can make eye contact” |
|  | P3 | “Will people think I'm weird for going ‘well this person's autistic’ when they don't meet the very traditional idea of it and I guess that's that sort of held true really” |
|  | P2 | "Psychiatry is particularly difficult because we seem to consider ourselves on top of the food chain... psychiatrists [are] really closed-minded about learning new things" |
|  | P4 | "Adult psychiatrists are kind of stuck with this idea of what autism is... they still think autism means this little white boy with their trains literally lining them up" |
|  | P2 | "They still seem to come out with that picture you know which I used to have in my textbook when I was studying, that boy lining up toys, that's autism, that's the what they expect to see - people lining up" |
|  | P7 | "Nobody actually quite knows how to deal with it... the majority don't necessarily have the training or experience to feel comfortable about it... where they are dealing with it they're seeing it as a disorder" |
|  | P5 | "They didn't know what to say because they've got no frame of reference... they couldn't say 'oh yes we've spotted it' because they don't know and they couldn't say 'oh don't think that's likely' because they don't know, so silence was produced" |
| **Theme 6:**<br>**Inaccessible**<br>**services** | P4 | "There are like a million of hoops you have to jump through to actually get to the assessment pathways and... a lot of people get screened out before any kind of in depth assessment takes place... I'm kind of thinking that this person should not have been screened out" |
|  | P2 | "I'm working in CAMHS and I'm seeing very obviously autistic women rejected by the mad process" |
|  | P2 | "GPs and psychiatrists just don't believe you... solutions available but only if you are believed in the first place" |
|  | P5 | "If I did not have the money to pay to go privately I would probably be at the point of self-identifying... if I wasn't somebody who could access articles and understood what they said I would be sort of left in a bit of a hole really" |
|  | P3 | "The waiting list on the NHS is so ridiculous... three years for something that somebody knows that she needs" |
|  | P4 | "If you can use the phone one day or not the other that's manipulation... pretending not to be able to talk on the phone today because two weeks ago you spoke to so-and-so" |
|  | P4 | "What we were doing in CAMHS... reasonable adjustments... people will be like 'oh this is just CAMHS mollycoddling you... we do it different in adults'... they strip it all and then... the wheels come off and there is a mega crisis and they will be on the phone... 'you said they were fine when you handed them over'" |
|  | P4 | "The onus is on the autistic person to find a solution to inaccessible services" |
|  | P5 | "How on earth can we expect autistic patients who have no insider experience of healthcare to navigate this system" |
|  | P7 | "There's a whole set of overlaps between eating disorders, gender identity, neurodevelopmental, which means that people sometimes feel a bit at sea dealing with it all" |
| **Theme 7:**<br>**Resource constraints** | P3 | "If you are going to have it as a disorder and a deficit that comes along with funding then there has to be some degree of gatekeeping... that somebody is actually dysfunctional enough for that, but that doesn't fit with the reality of how neurodiversity works does it?" |
|  | P5 | "People prioritized are... in danger of losing their job, difficulties with police and other services, forensic type things, relationships breaking down, difficulties with children... they are understandably prioritized" |
|  | P5 | "Psychiatrists are hugely overworked, there's not enough time, not enough resources, they need training, they need supervision as well" |
|  | P1 | "I'd tell people I'm autistic... except I'd be firmly told I wasn't... because I didn't have the [features], the intensity that was required" |
|  | P7 | "It's simply not been possible to be on top of my job for quite some time... it's now out of control... trying to... work through backlog... with 150 new patients waiting that's just not doable... normal for the sector is about 30" |
|  | P2 | "I feel very frustrated... because the diagnostic process is not with my trust... and they have a three year wait list" |
|  | P2 | "A disorder of the privileged because only the privileged will be able to afford to get diagnosed" |
|  | P4 | "We have those separate pathways... there's so much overlap... so much over work... you finish one pathway and then start up another... once you're observing the child you can... look for different things" |
| **Theme 8:**<br>**Underutilised resource** | P3 | "Autistic doctors being out... it's fantastic for society as a whole but particularly for trying to challenge some of these awful outdated ideas in so many of my colleagues" |
|  | P6 | "Colleagues would think this person is not going to understand patients or get it... little realising that probably masses more patients have underlying autism and they will get them better than a neurotypical person" |
|  | P4 | "I feel quite passionate about it and I will expend the energy gladly... that's probably quite a positive experience for children to actually have an adult that cares enough" |
|  | P5 | "There's absolutely nobody else who would diagnose her or see her... so I trained myself up" |
|  | P2 | "I wish we could find some way to raise... these concerns without being... I don't know... I mean... it's not a whistleblower level thing... something that will make us feel empowered enough to be able to say that this is not acceptable, something, some processes some pathways... [if] we do raise those concerns and people quickly get it [that] we are not at the same place so far as autism is concerned" |
| **Theme 9:**<br>**Pathways to change** | P5 | "We've got to start with training... autism is everybody's business... there's mandatory training coming... basically it's every psychiatrist's role to be able to recognise autism and consider diagnosis" |
|  | P1 | "Get psychiatry on board identifying neurodevelopmental disorder and work with that... rather than mailing it off to a specialist" |
|  | P2 | "Our textbooks are really out of touch... [change] textbooks first... that's the start" |
|  | P4 | "Some high hopes for the new ICD 11 because it seems to be a bit more modern diagnostic criteria... may help move the conversation along a bit" |
|  | P7 | "A full neurodevelopmental diagnostic service... a national pathway" |
|  | P4 | "A neurodevelopmental service that would look... in a very holistic way... what are the strengths, what areas of support do they need" |
|  | P2 | "The developments in the Royal College [of Psychiatrists] ... are encouraging... there will be a paradigm shift... borderline and treatment resistant depression... will actually change, but I'm thinking 10 years down the line" |

### Theme 1: Autistic-to-autistic rapport as a transformative relational phenomenon

Participants described a distinctive therapeutic rapport with autistic patients characterised by comfort, authenticity, and deep empathy. Mutual recognition of shared neurodivergence – tacit or explicit – facilitated trust, emotional safety, and rapid therapeutic engagement. Patients responded with relief at being genuinely understood, frequently after years of difficulty connecting with conventional psychiatric services.

> *“It’s a bit obvious to everybody even if you don’t say it… the way you talk and communicate and don’t look at each other… when everybody is comfortable… we all kind of stopped doing the masking and the usual social stuff*.*” (P4)*

Several participants described accumulating disproportionately large caseloads of autistic patients, referred by colleagues who had recognised their therapeutic effectiveness without understanding its basis:

> *“When they see some young person who might be [autistic] they quickly send to me… they don’t know that this is how I see myself but they know that when I talk to the young people I seem to be able to make a difference more quickly… they don’t know why*.*” (P2)*

This referral pattern – informal, intuitive, and operating below the level of explicit recognition – was consistent across specialties and settings. Participants described a relational dynamic qualitatively different from standard clinical encounters: one in which the usual demands of masking and social performance were suspended, and in which both parties could communicate more directly. Parents of autistic children were frequently described as baffled witnesses to these encounters, watching their child respond to being understood in ways they had not seen before. The rapport described was not experienced as technique or strategy but as an authentic expression of shared neurotype – a clinical asset that colleagues informally recognised but could not name.

### Theme 2: Benefit of recognition and diagnosis

Recognition of neurodivergence produced substantial and sometimes life-changing benefit for patients. Primary gains included improved self-understanding, development of a positive autistic identity, and enhanced capacity for self-advocacy – consistently identified as more therapeutically significant than any pharmacological or behavioural intervention. Considering long-standing difficulties through a neurodevelopmental lens allowed clinicians to reduce unnecessary medication, reframe distress that had previously been treated as treatment-resistant, and deploy creative diagnostic approaches that circumvented delays. One participant described using neurodiversity terminology rather than a formal diagnostic label to convey recognition to patients not yet formally assessed, producing immediate therapeutic benefit.

The impact of recognition extended across the lifespan. Participants described elderly patients approaching the end of life who found comfort in finally understanding themselves as autistic rather than as lifelong failures – comfort requiring no formal diagnosis, only being seen. One participant described a patient who had lived for decades without recognition:

> *“The woman was extremely grateful because she finally realised that she wasn’t broken… she was just autistic*.*” (P5)*

This reframing – from failed neurotypical to successful autistic – was identified as the core mechanism of benefit, with implications for mental health extending far beyond formal psychiatric treatment. For those reached early, clinical gains could be dramatic; suicidality and self-harm that had persisted for years resolved when a neurodevelopmental lens was applied for the first time. Recognition was not contingent on formal diagnosis: being genuinely understood was itself therapeutic.

### Theme 3: Early recognition and education as preventive factors

Participants identified early neurodevelopmental recognition and neurodiversity-affirmative approaches as protective against the development of mental health problems, with education emerging as the critical upstream environment. The disproportionate mental health burden autistic people carry was understood not as an inherent feature of autism but as the predictable consequence of environments that fail to accommodate neurodivergent needs – consequences that, with appropriate recognition and adjustment, may be substantially preventable.

In school settings, participants described a consistent pattern in which autistic pupils who appeared compliant and unproblematic were in reality exhausting their full cognitive and emotional resource in maintaining that compliance:

> *“Just because somebody is sitting there quietly and not causing a ruckus does not mean that they are thriving*.*” (P4)*

The energy consumed by behavioural conformity left nothing for learning or wellbeing, and mental health crises in adolescence or adulthood were described as the predictable and preventable downstream consequence of unaddressed need. Reasonable adjustments in school settings were rarely provided, despite participants’ persistent advocacy. The preventive lens extended to family systems.

> *“My husband had recurrent depression his whole life and now with more understanding he’s like ‘if only I could have been myself earlier’… he’s never been allowed to be true to his neurotype… it’s just presented as… recurrent depression… that’s my worry all along ‘cause it’s in the family on both sides*… *how can I avoid this happening in the kids?” (P3)*

Several participants described identifying unrecognised neurodivergence in parents as among the highest-impact interventions available – whole families thriving once parental needs were recognised and addressed. The trajectory from unrecognised neurodivergence in childhood through to entrenched psychiatric presentation in adulthood was consistently framed not as inevitable, but as a system failure.

### Theme 4: Iatrogenic harm from non-recognition and systemic pathways to misdiagnosis

Where neurodivergence was not recognised, participants described widespread and pervasive iatrogenic harm. Patients who had never been evaluated through a neurodevelopmental lens were frequently found to have accumulated years of ineffective or actively harmful treatment, including inappropriate behavioural strategies, polypharmacy for presumptive mood or psychotic disorders, and psychological therapies poorly adapted to autistic cognition. Long-term patients were consistently harder to help; harm had become entrenched and was difficult to reverse.

The most consistently described pathway to harm was misdiagnosis with emotionally unstable or borderline personality disorder (EUPD / BPD), particularly among young autistic women:

> *“One of the commonest things I see is young female patients with the EUPD diagnosis being generally made by locums on very little evidence, largely because they’re annoying and they certainly self-harm… and then actually you start asking the right questions and discover they’ve got autism*.*” (P7)*

This diagnostic substitution was not experienced as random error but as a systemic pattern with structural drivers. Unlike any other psychiatric diagnosis – which a clinician may make on clinical grounds – autism diagnosis in many UK commissioning areas requires referral to specialist neurodevelopmental services, with waiting lists commonly extending to two or three years. Psychiatrists are rarely trained in autism assessment and frequently lack confidence to diagnose; and where clinical suspicion exists, commissioning rules may prohibit a clinical diagnosis without specialist MDT involvement. The asymmetry is stark: a personality disorder diagnosis can be made in a single clinical encounter, while the pathway to an autism diagnosis may take years. Participants described this not as a matter of individual clinical convenience but as a perverse incentive embedded in the architecture of the system itself – one that actively steers clinicians away from neurodevelopmental formulations regardless of clinical evidence.

Patients who received a personality disorder diagnosis in place of autism were at risk of punitive behavioural management – described by one participant as adding layers of punishment to someone who was simply autistic and could not make themselves any different. Inpatient wards were described as containing large numbers of patients with EUPD diagnoses who had never been offered any evidence-based therapy, having been effectively blacklisted from treatment alongside the diagnosis itself.

Several participants suggested that for the majority of presentations, the specialist referral pathway is not clinically necessary: a psychiatrist with appropriate training should be able to make a clinical autism diagnosis on the basis of clinical assessment, as they would any other diagnosis. The requirement for specialist referral is experienced not as a quality safeguard but as an additional barrier with no equivalent in the rest of psychiatric practice.

### Theme 5: Knowledge gaps and stereotypes

Participants described pervasive and entrenched knowledge gaps about autism across mental health services. Reliance on outdated diagnostic stereotypes, particularly a stereotypically male, childhood-onset presentation, led to systematic failure to recognise autism in women, people from minoritised ethnic groups, those with strong language skills, and those who had developed effective masking strategies. Derogatory and dismissive attitudes towards autistic patients were reported as commonplace, including among senior and respected colleagues.

Multidisciplinary team members, particularly nursing staff, were perceived as more open to reconsidering formulations than consultant psychiatrists, who were described by several participants as particularly resistant to updating their clinical frameworks.

Participants who raised autism as a diagnostic possibility for patients in their services frequently encountered scepticism or dismissal:

> *“The people that make it to us are much more likely to be autistic than the general population, but that hasn’t filtered through to CMHT understanding at all yet… and so as a lone voice I look like I’m the one who’s got it wrong*.*” (P5)*

This epistemic dismissal, in which the clinician with the greatest expertise was presumed to be over-invested rather than correct, was felt to have direct consequences for patients, whose diagnostic formulations went unchanged and whose needs remained unmet. Participants described having to ration how frequently they raised autism as a possibility, for fear of being perceived as having a personal agenda rather than sound clinical judgement. Curricular reform, with training delivered by autistic people rather than parents or non-autistic professionals, was consistently identified as the foundational requirement for change.

### Theme 6: Inaccessible services

For many autistic people, barriers to recognition begin before they reach mental health services at all. Participants described gatekeeping at the level of primary care and referral triage as a significant source of missed diagnosis: General Practitioners frequently failed to recognise autistic presentations, particularly in women and those who had developed effective masking strategies, and referrals were not made. Formal screening tools used to triage referral requests were poorly sensitive to presentations outside the stereotypical profile, resulting in autistic people being excluded before reaching specialist assessment. One participant reflected that she herself would not have passed the screening criteria used by her own service:

> *“I think I would be missed… I don’t think I would have made it through to the autism team*.*” (P5)*

The irony was acute: the clinician potentially best placed to recognise autism in her patients may not have been identified as autistic by the pathway designed to find them.

Mental health services routinely failed to implement reasonable adjustments for autistic people, with consequences falling hardest on those in greatest need. Crisis provision was identified as a particularly stark example: access to most crisis services depends on telephone contact – a mode of communication that many autistic people find difficult in ordinary circumstances and may find impossible when most distressed:

> *“The prime example is the crisis teams… the only way you can get help is by phone, which immediately excludes most autistic people… I can obviously talk on the phone, but if I happen to be in a crisis and really struggling, there’s no way I could pick up the phone*.*” (P4)*

When autistic patients presented inconsistently – able to use services on some occasions but not others – this was routinely interpreted by clinical staff as manipulation or evidence of secondary gain, rather than as the natural variability of autistic experience under differing levels of stress. This framing generated clinical notes that interpreted autistic behaviour through a neurotypical lens, compounding diagnostic distortion across subsequent encounters.

Transitions between services, particularly from CAMHS to adult psychiatry, were identified as periods of predictable and preventable crisis. Reasonable adjustments established during childhood were routinely withdrawn on transfer, framed by adult services as inappropriate dependence; mental health crises reliably followed. Participants also highlighted the particular vulnerability of autistic people presenting to eating disorder and gender identity services, where co-occurring neurodivergence was frequently unrecognised, and to old age psychiatry, where autism was rarely considered as a diagnostic possibility at all.

### Theme 7: Resource constraints and diagnostic thresholds

Participants consistently identified a structural tension between the imperative to recognise autistic patients and the resource constraints governing what recognition can offer in practice. Assessment pathways are rationed: referrals are accepted only where functional impairment meets threshold criteria, and waiting lists for specialist neurodevelopmental assessment commonly extend to two or three years. This created a distinctive moral dilemma for clinicians with the insight to identify who needs assessment:

> *“You could then ask the GP to refer them but you know they won’t get seen for two or three years – is that unfair for a person having that question mark for two or three years when you’re not quite sure?” (P5)*

The threshold question itself was identified as a deeper structural problem at odds with neurodiversity-affirmative values. Recognising neurodivergence as natural human variation – the philosophical foundation of effective practice described in earlier themes – sits in direct tension with gatekeeping systems that require autism to constitute a disorder producing measurable functional harm before assessment is justified:

> *“This is actually the biggie in terms of resources, what is caseness in terms of what justifies the expenditure of resources on it… we cannot take curiosity referrals, if somebody is not needing the diagnosis made then we can’t afford to give them the time… and it’s not fair but I don’t know what to do about it, ‘cause we simply do not have resources to do it*.*” (P7)*

Those prioritised for assessment were consistently those in immediate crisis – facing loss of employment, relationships, or liberty – while those who would benefit most from early recognition were frequently excluded. This tension was experienced not as an abstract policy problem but as a daily clinical reality, which carried a cost in preventable harm.

### Theme 8: An underutilised resource

Participants expressed significant frustration at their limited capacity to improve patient outcomes beyond their individual clinical encounters. Possessing an insider understanding of what autistic patients need, and able to observe directly the harm caused by its absence in colleagues’ caseloads, they nonetheless faced persistent institutional and professional resistance to acting on that knowledge at a systemic level. This resistance had direct consequences for patients who remained under the care of clinicians without the relevant expertise, and whose needs went unmet or were actively mismanaged as a result.

The practical constraint was described with striking clarity:

> *“I have to almost ration how often I speak about autism in relation to patients, because if I bring it up too often I look like I’ve got a bee in my bonnet… because people don’t understand why I keep saying it, whereas the studies show that it’s very common in daily psychiatry practice*.*” (P5)*

The experience of being the most informed clinician in a system that interprets that expertise as bias was a consistent source of professional and moral distress. Several participants described the emotional toll of continuing to advocate for patients in the face of epistemic dismissal, knowing that disengagement – the self-protective option – would cost patients the one advocate best placed to understand their needs. The distinctive capacity autistic psychiatrists bring to patient care – rapid recognition, empathic rapport, and insider understanding of systemic failure – remains largely invisible to the profession, unrecognised in workforce planning, and therefore unavailable to the vast majority of autistic people who might benefit from it.

### Theme 9: Pathways to change

Despite the systemic failures described across preceding themes, participants articulated a clear and coherent vision of what change would require – grounded not in abstract aspiration but in clinical experience and insider understanding of precisely where and why the system was failing.

Training reform was identified as the essential starting point, with participants emphatic that it must be delivered by autistic people rather than parents or non-autistic professionals:

> *“I would really love to see more training that is actually delivered by autistic people… just wheel out some autistic people who can speak for themselves. Groundbreaking*.*” (P4)*

The goal was not specialist knowledge confined to neurodevelopmental teams but a fundamental shift in psychiatric culture; participants described autism as ‘everybody’s business,’ requiring every psychiatrist to be able to recognise it.

Structural reform of diagnostic services was felt to be equally urgent. Participants called for integrated neurodevelopmental services capable of assessing the full range of neurodevelopmental conditions holistically, replacing sequential single-category pathways with a national neurodevelopmental diagnostic infrastructure.

Underpinning all of this was a call for a fundamental reconceptualisation of neurodivergence itself – away from a deficit model and towards an understanding of autism as part of natural human variation:

> *“We have to see neurodivergence as just another thing like hair colour or height… not necessarily a disorder in itself, but… can predispose you towards certain things… if you have autism you’re more likely to have anxiety and OCD and chronic depression… that’s how it is… autism itself is just a difference… a different way of being*.*” (P7)*

The call to action extended beyond psychiatric services. Education and social care were identified as requiring the same neurodiversity-affirmative shift, addressing the upstream determinants of autistic mental health rather than waiting for preventable harm to accumulate. The ambition was explicitly preventive: a system in which autistic people are recognised and accommodated from the earliest opportunity.

## Discussion

This study examined the perspectives of seven senior autistic psychiatrists on mental healthcare for autistic people, drawing on dual insider positionality to illuminate both what effective care looks like and where – and why – the system fails. Across nine themes, participants described a coherent picture: distinctive therapeutic rapport and patient benefit when neurodivergence is recognised; preventable harm – structural, systemic, and deeply entrenched – when it is not; and a workforce whose expertise remains largely unrecognised and under-resourced.

The rapport described in Theme 1 offers a clinical articulation of the double empathy problem applied to therapeutic relationships.^12^ Where standard psychiatric encounters may be shaped by mutual misunderstanding between non-autistic clinicians and autistic patients, autistic–autistic clinical encounters appear to resolve this difficulty – producing the features of comfort, authenticity, and depth of empathy described here. This finding has implications beyond individual clinical relationships: it suggests that the double empathy problem is a clinically significant phenomenon with measurable effects on diagnostic accuracy, engagement, and outcomes. Notably, the same dynamic that produces therapeutic benefit in clinical encounters may suppress ADOS-2 item B-13, which scores reduced rapport as indicative of autism – the very rapport that helps autistic patients may obscure their diagnosis in standardised assessment contexts.^11,26^

The systemic misdiagnosis pathway described in Themes 4, 5 and 7 has structural rather than individual drivers. The diagnostic asymmetry we identify – in which any psychiatric diagnosis can be made clinically while autism diagnosis in many UK commissioning areas requires specialist referral with multi-year waiting lists – appears to create an architecturally embedded incentive to formulate autistic presentations as personality disorders. This is not primarily a problem of individual clinician knowledge or attitude, though both require attention, but of system design. Reform of diagnostic commissioning structures, including enabling psychiatrists to make clinical autism diagnoses without mandatory specialist referral, is a prerequisite for reducing the iatrogenic harm documented here. Empowering psychiatrists to diagnose autism clinically would have multiple compounding benefits: removing the structural incentive to formulate autistic presentations as personality disorders; reducing the years-long waits that currently defer recognition and allow harm to accumulate; addressing the diagnostic privilege gradient in which access to assessment depends on financial resources or professional connections; and utilising the clinical expertise that, as this study demonstrates, already exists within the psychiatric workforce.

The inaccessible service landscape described in Theme 6 reflects the same structural logic. Services that fail to provide reasonable adjustments, or routinely remove those previously established on transition to adult care, and clinical framings that interpret autistic variability as manipulation rather than neurological reality, are not isolated failures of implementation – they are the predictable output of systems designed by and for neurotypical users. Embedding reasonable adjustments as standard rather than exception is a prerequisite for equitable care.

The preventive argument in Theme 3 extends the analysis beyond mental health services. Participants identified education as the upstream environment in which the mental health burden of unrecognised autism accumulates, and family recognition as a high-impact intervention point. These findings align with growing evidence that mental health outcomes for autistic people are substantially determined by environmental factors rather than autism per se, and that neurodiversity-affirmative approaches in schools and families may be more effective prevention than any downstream psychiatric intervention.^15, 27–30^

The epistemic dismissal described in Theme 8 – in which clinicians with the greatest relevant expertise are presumed to be over-invested rather than correct – constitutes a form of epistemic injustice with direct patient consequences.^14,31^ Addressing it requires not only cultural change but structural recognition of autistic psychiatrists as a workforce resource: one with distinctive expertise that should inform training, supervision, workforce planning, and service development. Neurodiversity-affirmative approaches developed by autistic clinicians, including the Autistic SPACE Framework, provide a foundation for this shift in clinical culture.^32^

Critically, participants did not only document failure. The coherent change agenda articulated in Theme 9 – training delivered by autistic people, diagnostic pathway reform that removes structural incentives for misdiagnosis, and neurodiversity-affirmative approaches embedded from the earliest opportunity – carries particular epistemic weight precisely because it comes from clinicians who inhabit both sides of the system. These are not recommendations generated from the outside looking in, but from practitioners who have experienced autistic mental healthcare as both clinicians and, in many cases, as patients or potential patients themselves. The dual insider positionality that enables distinctive clinical effectiveness also enables a uniquely grounded account of what structural change would require, and what it would make possible.

### Strengths and limitations

The autistic-led design and insider methodology are strengths: dual insider positionality enabled participants to share perspectives likely inaccessible to non-autistic researchers, and the first author’s positionality as an autistic clinician facilitated interpretive depth.^19^ The sample is small and purposively drawn from ADI, limiting transferability to autistic psychiatrists not connected to autistic professional networks, or who have not yet recognised themselves as autistic. Participants were predominantly senior and UK-based; perspectives of trainees and those practising in different health system contexts may differ. The perspectives represented here arise from a specific subgroup of autistic people – senior autistic psychiatrists – and should not be assumed to represent the full heterogeneity of autistic experience, particularly for autistic people with intellectual disability, higher support needs, or limited verbal communication. These limitations are inherent to insider qualitative methodology and do not undermine the analytic contribution of the findings.

The majority of our participants self-identified as autistic without a formal diagnosis. Reasons for seeking or avoiding formal diagnosis will be reported elsewhere. Self-identification is a recognised practice within autistic communities, and our participants were psychiatrists whose clinical expertise includes autism diagnosis. Acceptance within the ADI Psych group, which consists of autistic and autism-diagnosing psychiatrists, provides community confirmation of autistic identity. We note that scrutiny of the data leaves us in no doubt as to the autistic identity of all participants.

## Conclusion

Autistic psychiatrists offer a uniquely informed perspective on mental healthcare for autistic people – one grounded in simultaneous membership of the clinical profession and the autistic community. Their accounts document both what effective care looks like and the systemic conditions that prevent it: inadequate training, structural diagnostic barriers, inaccessible services, and a workforce whose distinctive expertise remains largely unrecognised. The mental health burden autistic people carry is not an inevitable consequence of autism; it is substantially the product of systems not designed for them. The insights reported here point toward concrete change: curricular reform with training delivered by autistic people, diagnostic pathway redesign that removes structural incentives for misdiagnosis, neurodiversity-affirmative approaches embedded in education and healthcare from the earliest opportunity, and recognition of autistic psychiatrists as a clinical and epistemic resource.

## Data Availability

Data cannot be shared publicly due to ethical restrictions and risk of participant identification. Deidentified data may be available from the corresponding author on reasonable request, subject to appropriate ethical approvals.

## AI declaration

Claude (Sonnet 4.6, Anthropic, claude.ai) was used to assist with manuscript preparation, including literature search support and structural editing. All content was reviewed, verified, and approved by the authors, who take full responsibility for the accuracy and integrity of the published work.

## Author Contributions

Conceptualisation: MD, SCKS

Data analysis: MD, NC

Investigation: MD

Methodology: SCKS

Supervision: NM, EC, SCKS

Writing – original draft: MD

Writing – review & editing: MD, NC, NM, BG, EC, SCKS

